# Baseline inflammatory status affects the prognostic impact of statins in patients with peripheral arterial disease

**DOI:** 10.1101/2023.02.28.23286598

**Authors:** Kentaro Jujo, Daisuke Ueshima, Takuro Abe, Kensuke Shimazaki, Yo Fujimoto, Tomofumi Tanaka, Teppei Murata, Toru Miyazaki, Michiaki Matsumoto, Hideo Tokuyama, Tsukasa Shimura, Ryuichi Funada, Naotaka Murata, Michiaki Higashitani, Toma-Code Registry Investigators

**Affiliations:** Department of Cardiology, Nishiarai Heart Center Hospital, Tokyo, Japan; Department of Cardiology, Kameda Medical Center, Chiba, Japan; Department of Cardiology, Toranomon Hospital, Tokyo, Japan; Department of Cardiology, Sakakibara Heart Institute, Tokyo, Japan; Department of Cardiology, Tokyo Metropolitan Geriatric Medical Center, Tokyo, Japan; Department of Cardiology, Ome Municipal General Hospital, Tokyo, Japan; Department of Cardiology, Nihon University Hospital, Tokyo, Japan; Department of Cardiology, Kawaguchi Cardiovascular and Respiratory Hospital, Saitama, Japan; Department of Cardiology, Yokohama City Minato Red Cross Hospital, Kanagawa, Japan; Department of Vascular Medicine, Cardiovascular Hospital of Central Japan, Gumma, Japan; Department of Cardiology, Tokyo Medical University, Tokyo, Japan; Department of Cardiology, Tokyo Medical University Ibaraki Medical Center, Ibaraki, Japan, Ibaraki, Japan

**Keywords:** Peripheral artery disease, Endovascular therapy, C-reactive proteins, Statins

## Abstract

**Background:** Statins bring favorable effects on the clinical prognosis of patients with atherosclerotic disease partly through their anti-inflammatory properties. However, this effect has not been fully verified in patients with peripheral arterial disease (PAD). We aimed to test whether statins exert different prognostic effects depending on the degrees of inflammation in patients with PAD.

**Methods:** This study was a sub-analysis of a multicenter prospective cohort of 2,321 consecutive patients with PAD who received endovascular therapy (EVT). After excluding patients without information on C-reactive protein (CRP) levels at the time of index EVT, 1,974 patients (1,021 statin users and 953 non-users) were ultimately analyzed. Enrolled patients were classified into four groups depending on CRP levels: low CRP (<0.1 mg/dL), intermediate-low CRP (0.1–0.3 mg/dL), intermediate-high CRP (0.3–1.0 mg/dL), and high CRP (>1.0 mg/dL). A composite of death, stroke, myocardial infarction, and major amputation as the primary endpoint was compared between statin users and non-users in each CRP category.

**Results:** Overall, statin users showed a significantly lower event rate than non-users (log-rank, p<0.001). However, statin therapy was associated with significantly lower event rates only in the intermediate-high- and high-CRP categories (p=0.02 and p=0.008, respectively). Multivariable Cox regression analysis revealed that statin use was independently associated with the primary endpoint only in the high-CRP category (adjusted hazard ratio: 0.64 [95% confidence interval: 0.41–0.98]).

**Conclusion:** Statins may exert favorable prognostic effects in patients with PAD and highly elevated CRP levels but not in those with low to moderate CRP levels.

**Condensed abstract:** This multicenter retrospective study compared the prognostic effects of statins among patients with peripheral arterial disease (PAD) presenting diverse baseline C-reactive protein (CRP) levels [low CRP (<0.1 mg/dL), intermediate-low CRP (0.1–0.3 mg/dL), intermediate-high CRP (0.3–1.0 mg/dL), and high CRP (>1.0 mg/dL)]. Multivariable analysis showed that statin use was independently associated with a lower rate of death, stroke, myocardial infarction, and major amputation only in the high-CRP category. This suggests that statins may have favorable prognostic effects in patients with PAD and active inflammation.

## Introduction

Current treatment algorithms for the prevention of adverse cardiovascular events recommend statin therapy for patients with cardiovascular diseases and dyslipidemia. (1,2) Lipid-lowering therapy with statins is associated with a greater reduction in the incidence of major adverse cardiovascular events in patients with coronary artery disease (CAD). (3,4) However, half of all cases of myocardial infarctions and strokes occur among apparently healthy individuals with low-density lipoprotein (LDL) cholesterol levels below the currently recommended thresholds for treatment. (5) The progression of atherosclerosis is influenced by poor lipid profiles and chronic persistent inflammation. (6) This is attributed to the role of LDL-cholesterol as a major risk factor for atherosclerosis development and is inhibited by statins owing to their pleiotropic effects including lipid-lowering and anti-inflammation. (7) Inflammatory biomarkers, including (high-sensitivity) C-reactive protein (CRP) and serum interleukin (IL)-6, have been reported to be associated with an increased risk of cardiovascular events, independent of cholesterol levels. (8-11) The anti-inflammatory effect of statins prevents the progression of atherosclerosis in patients with CAD by reducing CRP levels. (11) However, to date, no prospective outcome trial has directly addressed the question of whether patients with LDL-cholesterol levels below the treatment target but with elevated CRP levels might benefit from statin therapy. (12,13)

Patients with peripheral arterial disease (PAD) generally have an advanced atherosclerotic status and present with comorbid CAD and other systemic atherosclerotic diseases. Despite appropriate medical therapy, supervised exercise therapy, and successful endovascular therapy (EVT), many studies have reported high risks of mortality and morbidity in patients with PAD with or without symptoms. (14) Statins reduce both LDL-cholesterol and CRP levels; however, it is difficult to determine the relative contribution of the reduction in each of these biomarkers to the observed clinical benefits. We aimed to address this knowledge gap by analyzing patients with PAD, divided into groups according to their CRP levels, who were registered in a prospective multicenter registry.

## Methods

### Study population and endpoints

This study is a post-hoc analysis of the Toma-Code Registry, which is a Japanese prospective cohort of 2,321 consecutive patients with PAD who were treated with EVT in 34 hospitals between August 2014 and August 2016. The study protocol was approved by the Ethics Committee at Sakakibara Heart Institute (reference no. 14-023) and the committees of each participating facility. This study was registered in the University Hospital Medical Information Network Clinical Trials Registry (UMIN-CTR No. UMIN000015100). The main findings of this registry have been previously published. (15) After the exclusion of patients without information on CRP levels at the time of index EVT and those who were lost to follow-up, 1,974 patients, including 1,021 statin users and 953 statin non-users, were ultimately analyzed in the current study. Referring to the prior report, (16) the patients were divided into four categories depending on the CRP level at the time of EVT: low CRP (<0.1 mg/dL), intermediate-low CRP (0.1–0.3 mg/dL), intermediate-high CRP (0.3–1.0 mg/dL), and high CRP (>1.0 mg/dL) (**Figure 1**). A composite of outcomes including death, stroke, myocardial infarction, and major amputation as the primary endpoint of this study was compared between statin users and non-users in each CRP category. This study was conducted according to the principles of the Declaration of Helsinki, and written informed consent was obtained from all patients before study entry.

**Figure 1.**
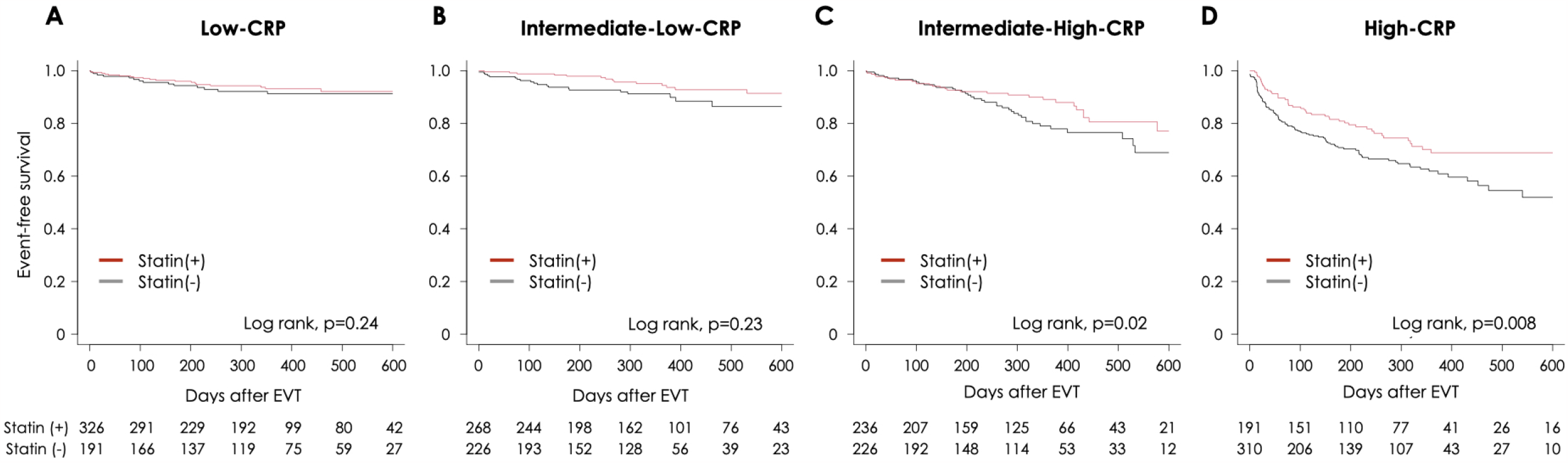
Study population. CRP = C-reactive protein; PAD = peripheral arterial disease.

### Data collection and follow-up

Demographic, laboratory, angiographic, and procedural data were collected from each patient’s hospital chart or database by independent researchers, according to predetermined definitions, and all data were collectively managed by the study office. Follow-up data were obtained from hospital charts or by contacting the patients or their family members via telephone. Patients were followed-up at 1, 6, 12, 18, and 24 months after the index EVT. We evaluated patients’ baseline characteristics, including age; sex; body mass index (BMI); history of smoking; history of revascularization therapy and lower limb amputation; having other comorbidities, including CAD, cerebrovascular disease (CVD), chronic kidney disease (CKD); complete blood count [white blood cell (WBC) and platelets]; hemoglobin level; blood chemistry test results [albumin, creatine phosphokinase, and CRP levels and lipid profiles including total-, LDL-, and high-density lipoprotein (HDL)-cholesterol levels]; and clinical presentations of PAD (Rutherford classification and ankle brachial pressure index of the target limb) at the time of EVT. The prescription of medications including aspirin, P2Y12 inhibitors, cilostazol, oral anticoagulants, beta-blockers, angiotensin-converting enzyme inhibitors or angiotensin II receptor blockers, calcium channel blockers, and statins at discharge was recorded. CKD was defined as an estimated glomerular filtration rate (eGFR) <60 mL/min/1.73 m^2^ and urine albumin level >30 mg/g. The location and distribution of the target lesions of EVT were also recorded: iliac, femoropopliteal, below-the-knee (BTK), graft vessel, and unilateral/bilateral.

### Statistical analysis

Values are expressed as the mean ± standard deviation, median and interquartile range, and percentiles, as appropriate. The independent Student’s t-test and non-parametric equivalent Mann– Whitney U test were used to compare statin users and non-users with respect to continuous variables. The Fisher’s exact test was used to evaluate categorical variables. The chi-square test was used to compare the Rutherford classification between statin users and non-users. A composite of outcomes including death, stroke, myocardial infarction, and major amputation was evaluated using Kaplan– Meier curves, differences among the four subgroups were assessed using the log-rank trend test, and differences between statin users and non-users were assessed using the log-rank test. Univariate Cox hazard regression analysis was performed to evaluate the association between statin use and prognosis after EVT. Variables reaching a level of significance of p <0.1 in the univariate analysis and considered clinically significant were included in the multivariable model.. Multivariable Cox hazard regression analysis was performed to exclude confounding factors and identify independent risk factors for the primary composite endpoint. To evaluate the hazard ratio (HR) of the primary endpoint based on statin use in the intermediate-high-CRP category, variables such as age, sex, BMI, smoking history, dyslipidemia, CKD, aortic stenosis, serum albumin level, P2Y12 inhibitor use, iliac lesion, BTK lesion, and stent implantation were used to adjust the multivariable model. For the high-CRP category, age; sex; BMI; hypertension; CKD; heart failure; CAD; CVD; hemoglobin, albumin, and LDL-cholesterol levels; bilateral target; BTK lesion; stent implantation; and procedural complications were used. Two-tailed P-values <0.05 were considered statistically significant. Statistical analyses were performed using R software, version 3.3.0 (R Foundation for Statistical Computing, Vienna, Austria).

## Results

Of the 2,321 consecutive patients with PAD who underwent EVT, 282 patients lacking CRP data and 65 patients lost to follow-up were excluded (**Figure 1**). Among the enrolled 1,974 patients, 517 (26.2%) were assigned to the low-CRP category, 494 (25.0%) to the intermediate-low-CRP category, 462 (23.4%) to the intermediate-high-CRP category, and 501 (25.4%) to the high-CRP category. In addition, at the time of EVT, 1,021 (51.7%) patients received statins, whereas 953 (48.3%) patients did not. A total of 326 (63.1%) patients in the low-CRP category, 268 (54.3%) in the intermediate-low-CRP category, 236 (51.1%) in the intermediate-high-CRP category, and 191 (38.1%) in the high-CRP category received statins.

### Patient profiles

Baseline patient profiles are presented in **Table 1**. In the whole cohort, the average age was 73 years, 72% of the patients were males, and statin users were significantly older and more obese. Diverse comorbidities, such as hypertension, CKD, hemodialysis, and CAD, were significantly more prevalent among statin users. The statin user group included fewer patients with atrial fibrillation or the on-ambulant status but frequently received prior EVT. In terms of laboratory data, statin users had higher hemoglobin levels and lower WBC, platelet, and CRP levels. Overall, statin users received more intensive medical treatment, except with cilostazol and oral anticoagulants. Angiography revealed that statin users were likely to have fewer BTK lesions, be in lower Rutherford classes, and receive more stent implantations.

**Table 1.**
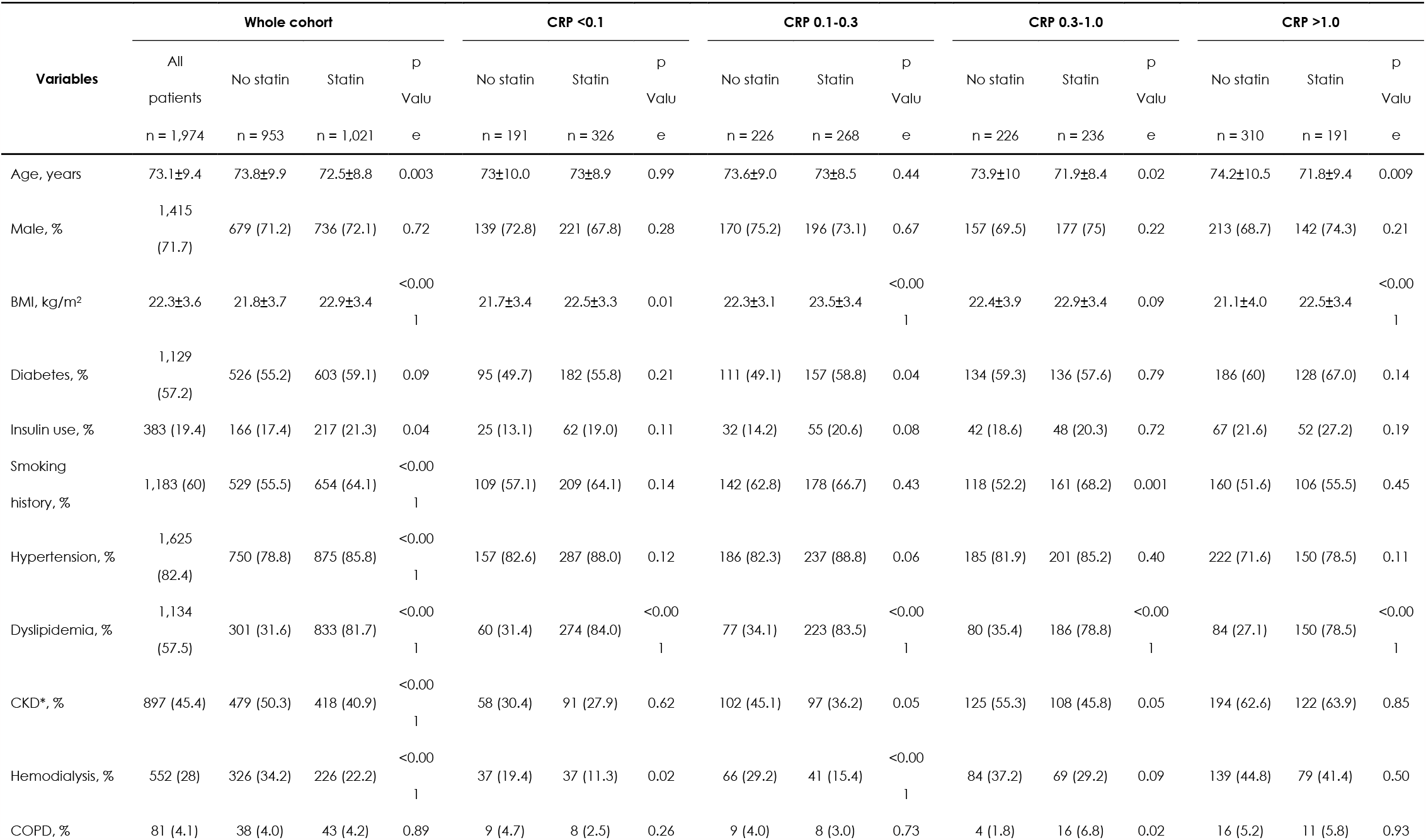
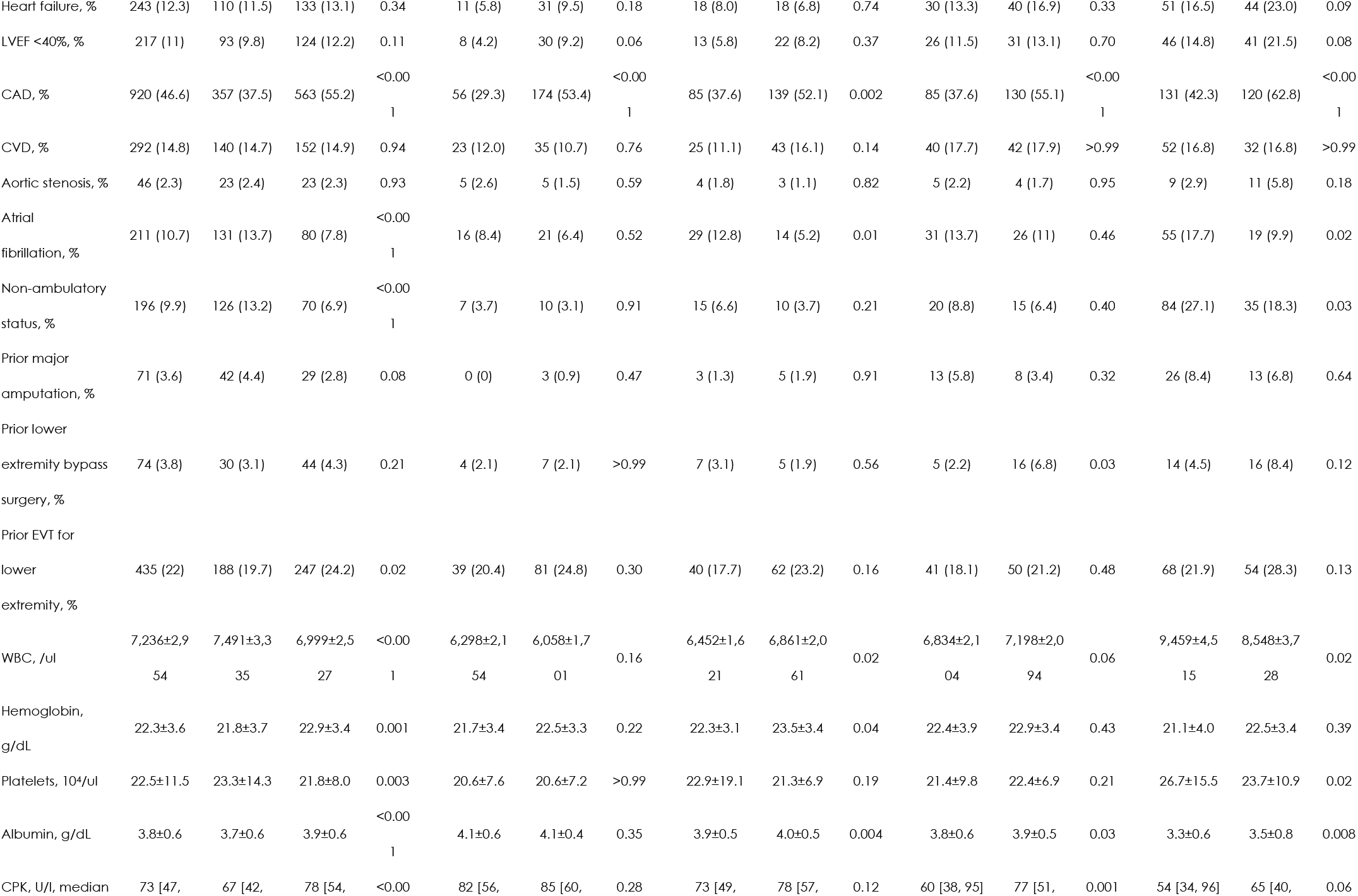
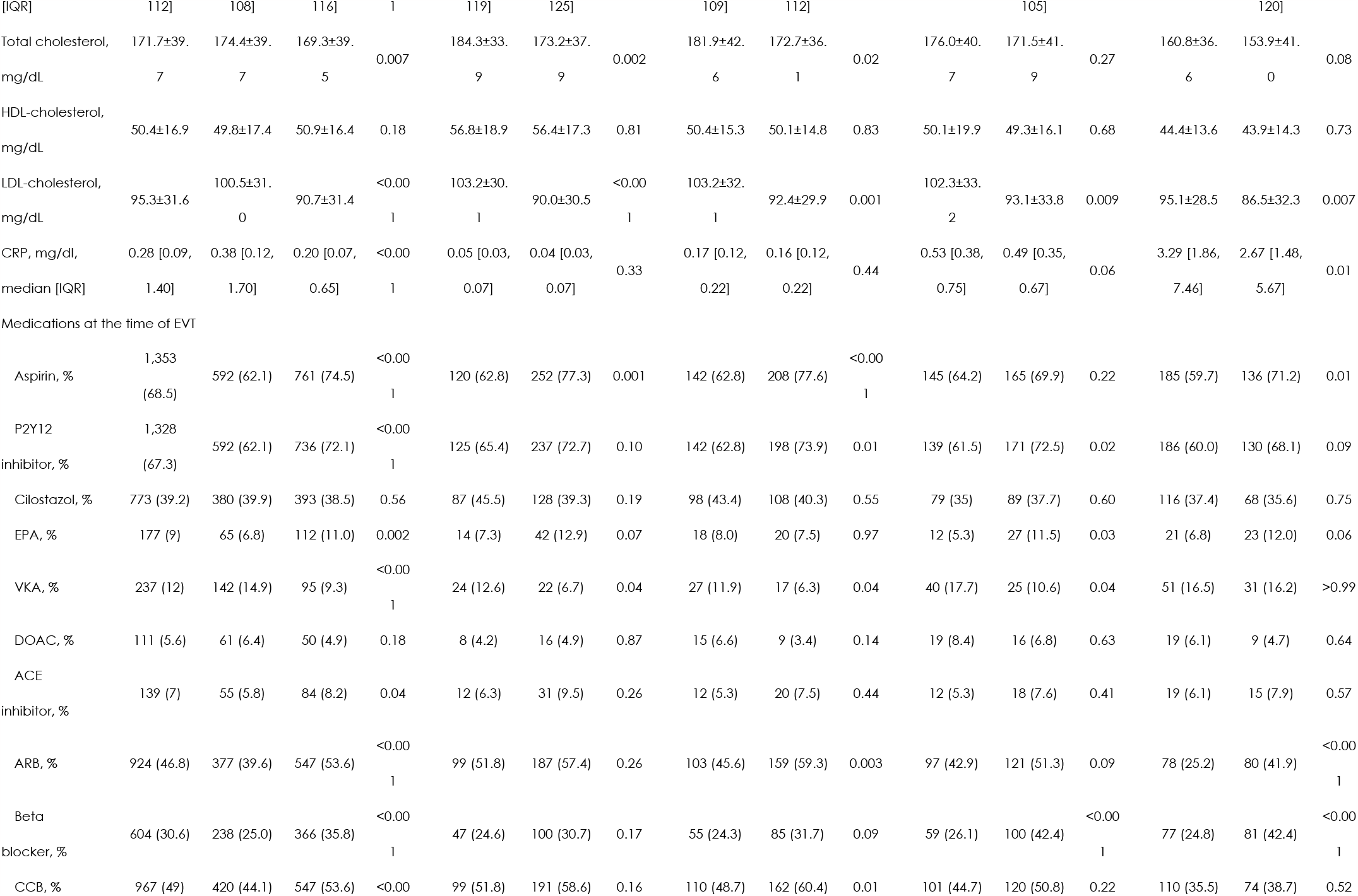
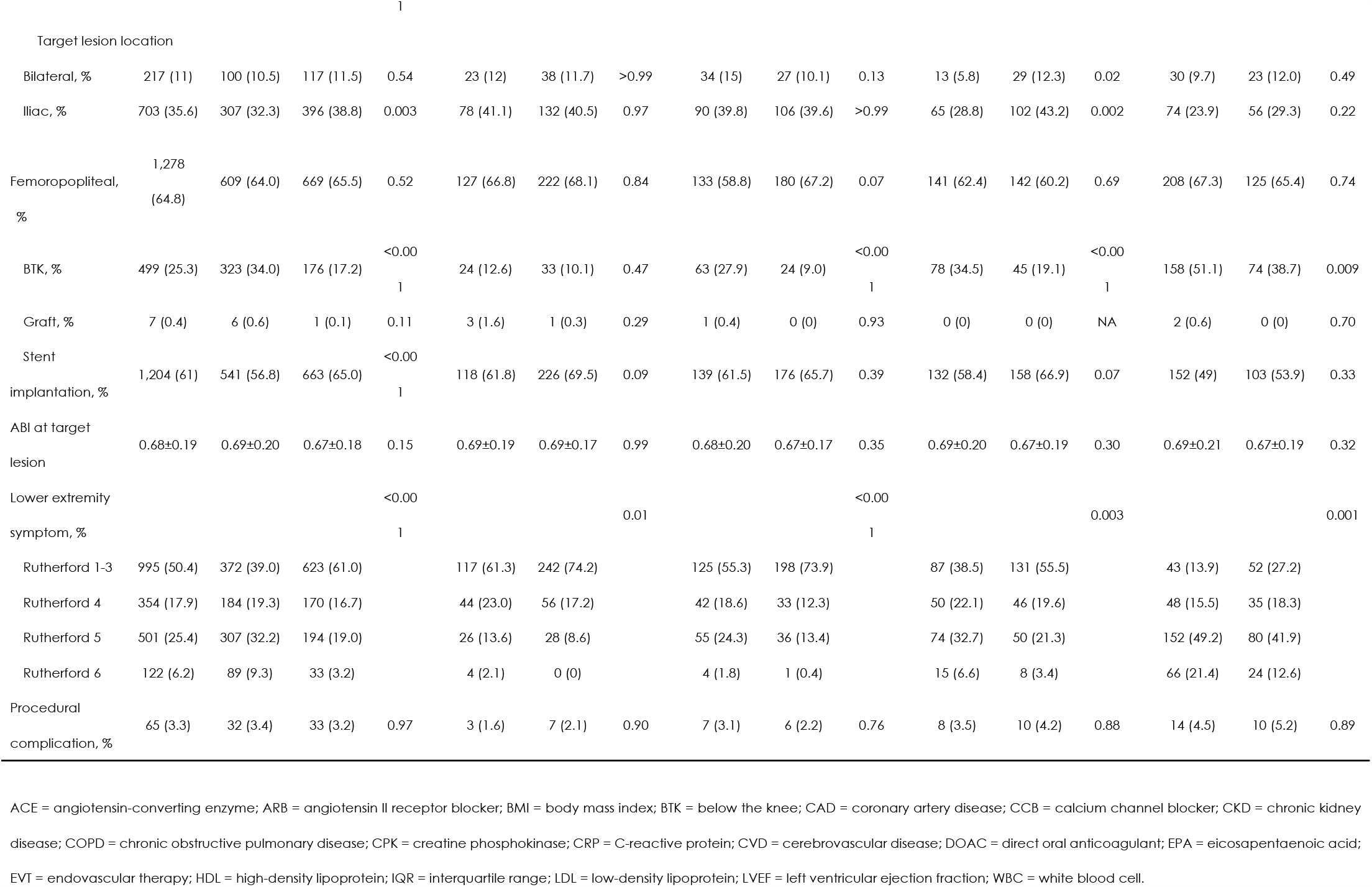
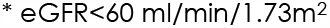
Baseline patient profiles

In each CRP category, statin users were significantly younger in the higher CRP categories, and the results were consistent with those of the entire cohort. The statin user group included more patients undergoing regular hemodialysis only in the lower CRP categories. Patients in the higher CRP categories had higher Rutherford classes, and the statin user group included more non-ambulatory patients only in the high-CRP category. Baseline CRP levels were significantly lower in statin users than that in non-users, only in the high-CRP category.

### Clinical outcomes

Overall, during the observation period [median 316 (interquartile range: 177–411) days), statin users had a significantly lower event rate of the primary endpoint than non-users (log-rank: p < 0.001) (**Figure 2A**). While patients in the lower CRP categories showed a lower event rate (log-rank test for trend: p < 0.001) (**Figure 2B**).

**Figure 2.**
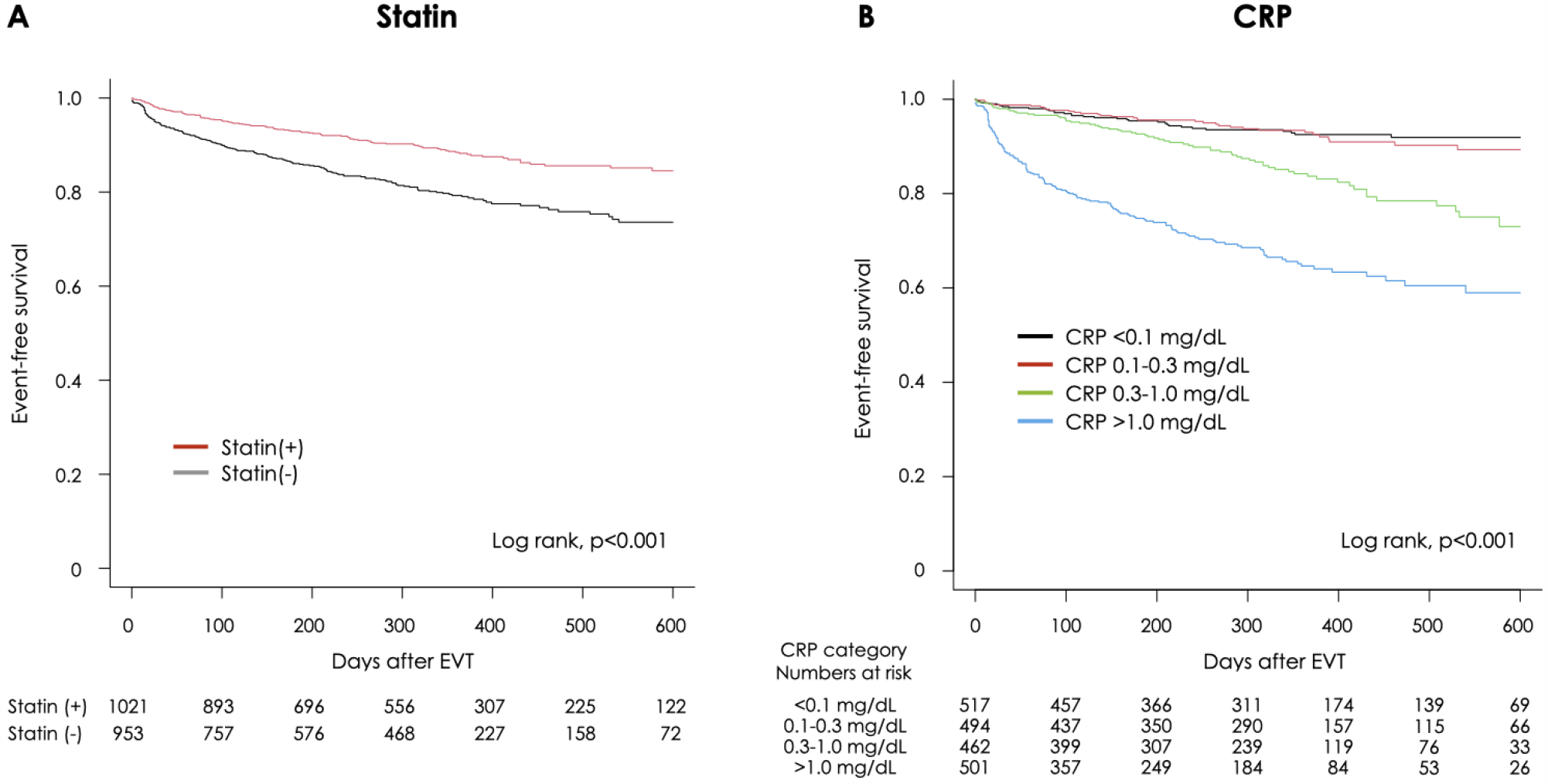
Incidence of the primary endpoint regarding statin use and CRP levels. Kaplan–Meier curves for freedom from a composite of outcomes including death, stroke, myocardial infarction, and major limb amputation in patients who were treated with and without statins (A) and in those who were categorized into four different groups according to CRP levels (B). CRP, C-reactive protein; EVT, endovascular therapy; pts, patients; inter-low, intermediate-low; inter-high, intermediate-high.

**Figure 3** shows the Kaplan–Meier curves between statin users and non-users in the four different CRP categories when comparing the primary endpoint. In the intermediate-high-CRP and high-CRP categories, statin users had significantly lower rates of the primary endpoint (log-rank, p = 0.02, p = 0.008, respectively). However, in the other lower CRP categories, there were no statistically significant differences between statin users and non-users. Univariate Cox regression analysis in the high-CRP category showed that BMI, CKD, CVD, albumin, alanine aminotransferase, and statin use were associated with the incidence of the primary endpoint (**Table S1**). The multivariable model for patients in the high-CRP category revealed that statin use was still independently correlated with the primary endpoint, even after the adjustment of covariates [HR: 0.64, 95% confidence interval (CI): 0.41–0.98; **Table 2**]. For the intermediate-high-CRP category, statin use was not associated with the primary endpoint (HR: 0.65, 95% CI: 0.35–1.21).

**Table 2.** Hazard ratio of primary endpoint by statin use

| CRP category | Unadjusted |  |  | Adjusted |  |  |
| --- | --- | --- | --- | --- | --- | --- |
|  | HR | 95% CI | p Value | HR | 95% CI | p Value |
| CRP <0.1 | 0.61 | 0.26-1.40 | 0.24 |  |  |  |
| CRP 0.1-0.3 | 0.64 | 0.30-1.34 | 0.23 |  |  |  |
| CRP 0.3-1.0* | 0.54 | 0.32-0.91 | 0.02 | 0.65 | 0.35-1.21 | 0.18 |
| CRP >1.0** | 0.62 | 0.44-0.89 | 0.009 | 0.64 | 0.41-0.98 | 0.046 |

\* adjusted by age, sex, BMI, smoking history, dyslipidemia, CKD, AS, albumin, P2Y12, iliac lesion, BTK lesion, stent implantation
\*\* adjusted by age, sex, BMI, hypertension, CKD, heart failure, CAD, CVD, hemoglobin, albumin, AST, bilateral target, BTK lesion, stent implantation, procedural complication

**Figure 3.**
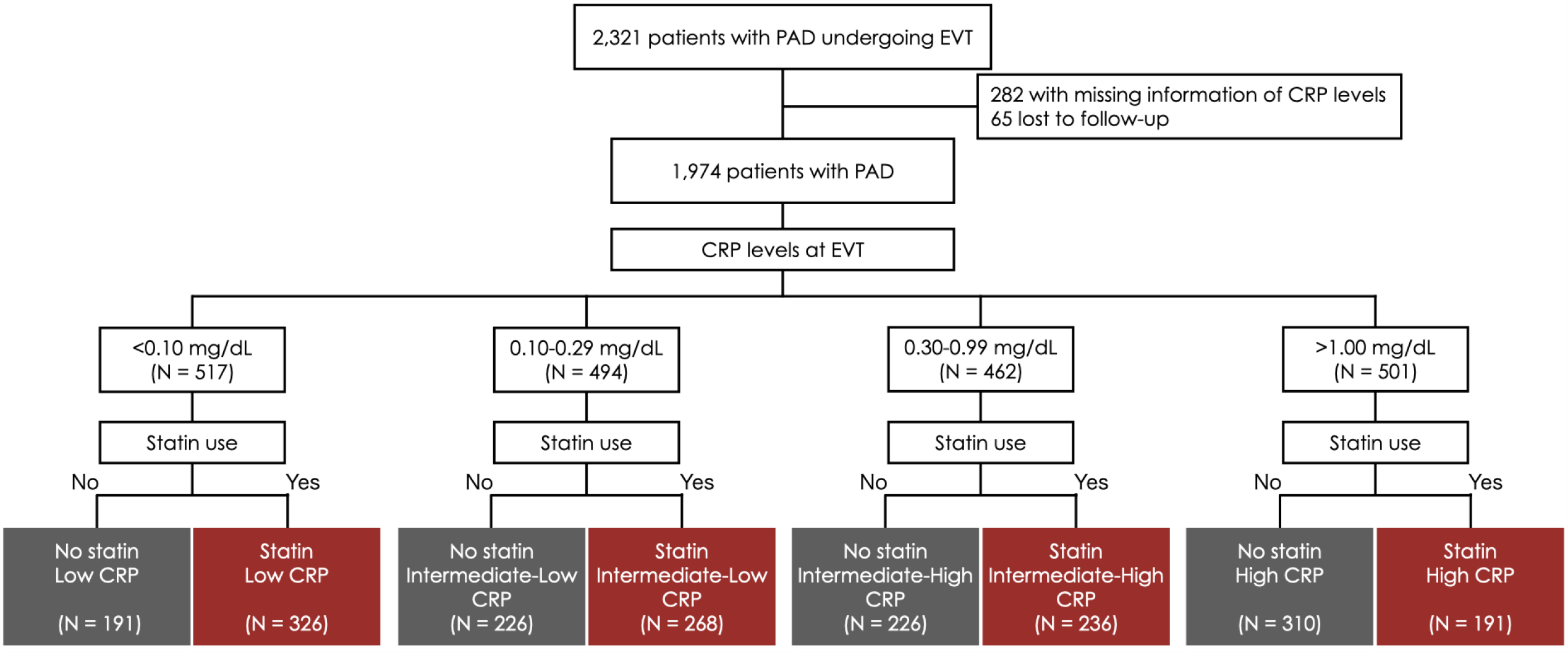
Comparison of the primary endpoint between statin users and non-users. Incidence of composite of outcomes including death, stroke, myocardial infarction, and major limb amputation in statin users (35) and non-users (gray) in the low-CRP (A), intermediate-low-CRP (B), intermediate-high-CRP (C), and high-CRP (D) categories. CRP, C-reactive protein; EVT, endovascular therapy; pts, patient.

The components of the combined endpoints were examined individually. Multivariable analysis showed that statin use was independently associated with the incidence of death from any cause (HR: 0.57; 95% CI: 0.40–0.82), only in the entire cohort. In any other category in any component, statin users did not show prognostic superiority compared to statin non-users (**Table S2**).

## Discussion

The principal finding of this study was that statin treatment did not affect the prognosis of patients with low to moderate CRP levels but was associated with improved prognosis in patients with CRP levels greater than 1.0 mg/dL at the time of EVT. We previously reported the prognostic benefit of statin use only in patients with PAD and elevated baseline CRP levels in a single-center database. (17) The present study validated this association using data from a large-scale multicenter prospective registry.

Prior placebo-controlled trials and meta-analyses evaluating the relationship between statin use and prognosis have revealed its benefits for all-cause and cardiovascular mortality. (18-24) These accumulated research results have proven the concept of the lower the better of lipid profiles. In contrast, no reduction in long-term cardiovascular mortality was observed in a trial of moderate-vs. high-intensity statin therapy in patients with prior myocardial infarction. (25) Most of the study population was in a stable condition; hence, the margin for the lipid-lowering therapy to work was low. Additionally, the JUPITER trial demonstrated the benefits of rosuvastatin treatment for cardiovascular outcomes of patients with high plasma CRP levels (>2.0 mg/dL) but without high LDL-cholesterol levels. (26) Collectively, sufficient experimental and clinical evidence has established statin pleiotropy in patients with CAD.

As for patients with PAD, there is limited evidence from randomized clinical trials on the benefits of statin treatment. In a large observational study that included low-risk patients with PAD, statin therapy was associated with a reduction in the incidence of major adverse cardiac events and all-cause mortality. (27) Consistent with the findings of this report, in the entire cohort of the current study, statin administration at the time of EVT was associated with improved long-term prognosis, and the baseline CRP levels well stratified the risk of adverse clinical outcomes. From this point of view, only patients with PAD having CRP levels greater than 1.0 mg/dL as the population with higher event rates achieved a statistical difference in the study endpoint by statin use. In this high-risk population, the distance between the Kaplan–Meier curves in the two groups began to widen very early after the index EVT. PAD can be regarded as a systemic, chronic inflammatory disorder with a direct relationship between the level of inflammation and its severity. (28) Prior reports have indicated that statins exert rapid effects before any lipid-lowering takes place and improve nitric oxide vascular bioavailability by increasing tetrahydrobiopterin synthesis and lowering the activity of NADPH oxidases in the vascular wall. (12,13) This also supported that the favorable prognostic effect of statins was predominantly through anti-inflammatory properties. Downstream inflammatory signaling by CRP and upstream signaling by IL-1b and IL-6 are involved in atherosclerosis-related systemic low-grade inflammation. The signaling mediators involved in atherosclerotic plaques and vascular wall inflammation have also been well characterized. (29)

Multiple mechanisms may contribute to improving the prognosis of patients with atherosclerosis; however, the present study indicates the potential of statins to work predominantly through the anti-inflammatory axis in patients with PAD, including the population with advanced atherosclerosis. Alternatively, the results may indicate that the favorable prognostic effects of statins are observed only in patients with “activated” vascular inflammation and “progressive” atherosclerosis. The role of inflammation in atherosclerosis is now well established, not only by studies measuring pro-inflammatory biomarkers in human plasma, (30) but also by Mendelian randomization studies revealing a causal relationship between genetic variation in cytokine signaling, such as the IL-6 receptor (31), and the risk of development of atherosclerotic disease phenotypes. Statins have a variety of pleiotropic properties, including the ability to induce dose-dependent decreases in the levels of CRP and other inflammatory biomarkers. (32) There are findings supporting the anti-inflammatory effects of statins, including the reduction of CRP levels in patients taking statins regardless of the level of decrease in LDL-cholesterol level. These anti-inflammatory effects are of particular importance in preventing the formation of atherosclerotic plaques and in preserving the functions of endothelial cells. (33) The present study showed that higher CRP levels were associated with worse clinical prognoses. Interestingly, in the current study, patients in the higher CRP categories received lower chances of statin treatment.

The legacy effects of statin treatment have been reported in a prior meta-analysis that the induction of statin itself was associated with prolonged improved outcomes in the remote phase after induction. (34) In the current study, we observed the relationship between statin use at the time of index EVT and subsequent clinical outcomes; however, we did not consider new induction of statins during the observation period. Given the proven legacy effect of statins, the findings of the current study indicate that earlier statin use may improve long-term outcomes in high-risk patients. Additionally, in the real clinical settings, there are few opportunities of intervention to reduce inflammation in patients with advanced and progressive atherosclerosis who have suspended inflammation. The results from the present study may help doctors select the appropriate timing and patients with PAD to receive the most benefit from statin treatment.

### Limitations

This study had some limitations. First, the study patients who underwent EVT and CRP testing were selected; therefore, a selection bias may have occurred. There is no evidence in this study that statins were newly prescribed during the index hospitalization or were already prescribed before hospitalization. In addition, the discontinuation or new induction of statins during the observation period was not considered in this study. Information on the types and doses of statins administered to patients was not collected; therefore, we could not determine the kind or dose-dependent effect of statins on prognosis. No therapeutic guidelines were pre-specified in the current study, and the therapeutic regime was at the discretion of the attending doctors in each hospital; therefore, some patients were indicated to receive lipid-lowering therapy with strong statins but did not receive the appropriate guideline-directed therapy. Details regarding the etiology of increased CRP levels were not examined. High-CRP levels may represent active infections, malignancies, or chronic inflammatory diseases, including collagen diseases, and the effects of statins on these conditions are unknown. CRP, but not high-sensitivity CRP, was set as the biomarker for inflammatory status in this study because high-sensitivity CRP was not routinely examined in all patients with PAD who were scheduled to receive EVT in each hospital. No changes in lipid profiles or inflammatory status were observed after discharge. Therefore, it is difficult to conclude which factor, between the anti-inflammatory and lipid-lowering properties of statins, plays a more important role in decreasing the rate of adverse clinical outcomes. Prospective statin entry studies that include CRP-categorized patients are required to clarify the prognostic impact of the anti-inflammatory effects of statins.

## Conclusions

This study evaluated the prognostic effects of statin therapy with respect to the degree of inflammation and found that statin therapy at the time of EVT was associated with improved clinical outcomes in patients with PAD and highly elevated CRP levels but not in those with low to moderate CRP levels. The initiation of statin therapy after EVT should be considered if patients show an activated inflammatory status.

## Data Availability

All data generated or analyzed during this study are included in this article. Further enquiries can be directed to the corresponding author.

## Acknowledgement

We would like to thank Editage (www.editage.jp) for English language editing.

## Abbreviations

CAD: coronary artery disease
CRP: C-reactive protein
EVT: endovascular therapy
PAD: peripheral artery disease

